# A New Approach for Timely Systematic Reviews of Rapidly Evolving Research: Identifying Evidence in Which We Can Place Our Confidence

**DOI:** 10.1101/2021.10.19.21265139

**Authors:** Jiayi Tong, Rebecca A. Hubbard, M. Elle Saine, Hua Xu, Xu Zuo, Stephen E. Kimmel, Adam Cuker, Yong Chen

## Abstract

Coronavirus disease of 2019 (COVID-19) has impacted the world in unprecedented ways since first emerging in December 2019. In the last two years, the scientific community has made an enormous effort to understand COVID-19 and potential interventions. As of June 15, 2021, there were more than 140,000 COVID-19 focused manuscripts on *PubMed* and preprint servers, such as *medRxiv* and *BioRxiv*. Preprints, which constitute more than 15% of all manuscripts, may contain more up-to-date research findings compared to published papers, due to the sometimes lengthy timeline between manuscript submission and publication. Including preprints in systematic reviews and meta-analyses thus has the potential to improve the timeliness of reviews. However, there is no clear guideline on whether preprints should be included in systematic reviews and meta-analyses.

Using a prototypical example of a rapid systematic review examining the comparative effectiveness of COVID-19 therapeutics, we propose including all preprints in the systematic review by assigning them a weight we term the “confidence score”. Motivated by our observation that, unlike the traditional journal submission process which is unobserved, the timeline from submission to publication for a preprint can be observed and can be modeled as a time-to-event outcome. This observation provides a unique opportunity to model and quantify the probability that a preprint will be published, which can be used as a confidence score to weight preprints in systematic reviews and meta-analyses.

To obtain the confidence score, we propose a novel survival cure model, which incorporates both the time from posting to publication for a preprint, and key characteristics of the study described in the content of the preprint. Using meta data from 158 preprints on evaluating therapeutic options for COVID-19 posted through 09/03/2020, we demonstrate the utility of the confidence score in weighting of preprints in a systematic review. Our proposed method has the potential to advance timely systematic reviews of the evidence examining COVID-19 and other clinical conditions with rapidly evolving evidence bases by providing an approach for inclusion of unpublished manuscripts.

## INTRODUCTION

The global pandemic of novel coronavirus disease 2019 (COVID-19) caused by severe acute respiratory syndrome coronavirus 2 (SARS-CoV-2) was first described in Wuhan, China in December 2019, and has since spread rapidly worldwide^1^. The symptoms of COVID-19 infection vary in severity from asymptomatic disease to pneumonia and life-threatening complications, such as acute respiratory distress syndrome, multisystem organ failure, and death. As of June 15, 2021, there have been more than 176 million reported cases, with over 3.8 million deaths in more than 200 countries^2^. To date, despite many COVID-19 symptomatic patients having been administered off-label and compassionate-use drugs, such as hydroxychloroquine, lopinavir-ritonavir, favipiravir, remdesivir, ribavirin, convalescent plasma, anti-IL-6 inhibitors, among others, the relative efficacy and safety of these drugs are still under investigation as increasing data emerges^3^. There is a critical need to conduct *timely and unbiased evidence synthesis* based on evidence from existing data.

As of June 15, 2021, there are more than 140,000 manuscripts on COVID-19 published or posted at PubMed, BioRxiv, and MedRxiv on COVID-19 from researchers all over the world^4,5^. These manuscripts cover a wide spectrum of important topics that can help us to understand the critical aspects of clinical and public health impacts of COVID-19, including the disease mechanism, diagnosis, treatment, and prevention as well as viral infection, replication, pathogenesis, transmission, host-range, and virulence. However, in addition to the evidence-based information, a vast amount of misinformation and rumours were immediately disseminated through social media outlets such as Twitter, Facebook, WhatsApp, Instagram, and Wechat^6,7^. The amount of information is increasingly overwhelming for clinicians, policymakers, researchers, and other stakeholders to process and appraise, and the widespread sharing of the information without being thoroughly vetted would be dangerous. “*We’re not just fighting a pandemic; we’re fighting an infodemic*,” said Tedros Adhanom Ghebreyesus, WHO’s director-general. Therefore, accurate, professional, and reliable information is in desperate need in this pandemic. A systematic review, which is a type of literature review that uses systematic methods to collect secondary data and critically appraise research studies, can be useful in summarizing the existing evidence of COVID-19 related research findings. In particular, meta-analysis plays a central role in quantitatively synthesizing evidence from multiple scientific studies which address related questions to obtain more reliable and generalizable conclusions.

A major consideration when performing a meta-analysis of COVID-19 related manuscripts is the large number of preprint articles on servers such as the BioRxiv and MedRxiv. These repositories facilitate the rapid dissemination of findings and are particularly suited to support efforts in understanding the disease in real-time as the outbreak unfolds and finding timely solutions. In early June, preprints accounted for more than 15% of COVID-19-related papers since January 2020, as compared to little more than 2% of all biomedical literature in the previous year (Editorial, 2020). However, these studies are not peer-reviewed, rushed to post, and highly heterogeneous in quality^8,9^. Such heterogeneity needs to be accounted for in the systematic reviews and meta-analyses, where studies with higher quality ought to be given more weight compared to studies with relatively lower quality. The lack of effective treatment for COVID-19 treatment has required providers to predominantly rely on the rapidly emerging and evolving body of literature regarding investigational therapeutics. Studies of poor methodologic quality which inform clinical practice guidelines risk undue harm to patients. The process of publication serves as a natural way to filter out studies with higher quality. Among the over 25,000 COVID-19 studies^5^ submitted to online preprint servers, it is reasonable to believe that only a portion of these studies, particularly those of higher quality, will ultimately be published. Ideally, if we know which study will be published in the end, we can assign more weight to those studies compared to those that will not be published. However, as the process of publication usually takes some time, most of the studies posted on the preprint servers are still under review, and the reader does not know whether they will be published in a peer-reviewed journal.

Motivated by the current rapid dissemination of studies related to COVID-19 through both peer-reviewed and non-peer-reviewed means, we aim to use statistical models to predict whether a study will be published. The probability of a study being published by a journal, which we term as the publication confidence score (CS), could be incorporated as an imputation weight in a meta-analysis. Since we can observe the time when a manuscript is published by the preprint server and the time when a study is published by a peer-reviewed journal, we hope to take the temporal information into consideration by using a survival modeling approach. If we treat the publication in a peer-reviewed journal as an event, it is natural to think that studies that will never be published as “cure”. We then model the process of publication using a survival cure model^10–12^, which predicts the probability of being “cured” (never being published in a journal) as well as the time to the event (being published). Factors such as sample size, whether the study is a randomized control trial (RCT), the number of citations that can be used to predict the “cure” can be extracted from the website using texting mining tools. Incorporating the published studies that are not submitted to any preprint servers, we can conduct weighted meta-analyses to synthesize the evidence.

## DATA AND METHOD

In **Figure 1** we present a Venn diagram comprising manuscripts that are submitted for public dissemination. Manuscripts that are posted to preprint servers such as BioRxiv and MedRxiv may also be submitted to journals for consideration for publication. On the other hand, manuscripts may be directly submitted to journals without posting to a preprint server, and some preprints are never submitted to journals for review. Thus far the majority of COVID-19 related papers in the public domain have been submitted directly to peer-reviewed journals, though this may be subject to change as the number of COVID-19 preprints continues to grow.

**Figure 1.**
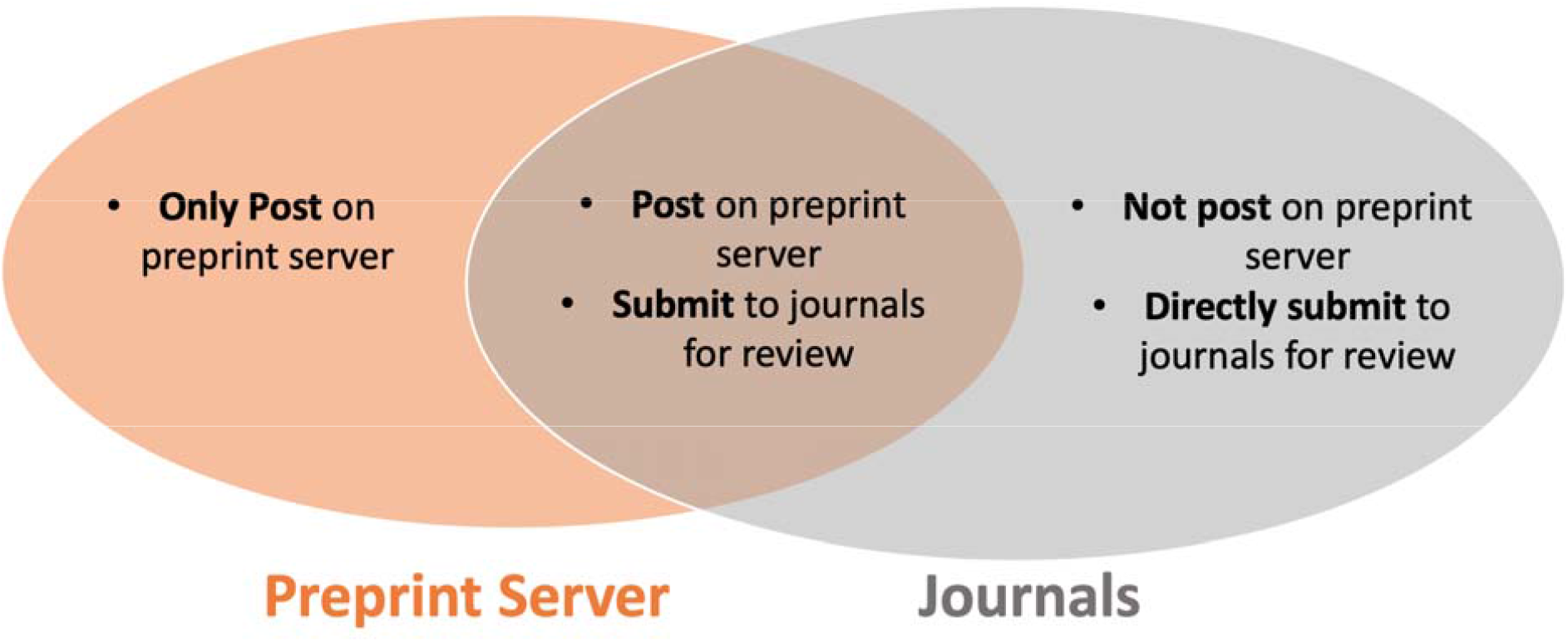
Venn diagram of manuscripts that enter the pipeline for public release

### Data

#### Databases and Search Strategy

##### Peer-reviewed

Infectious disease experts, including clinicians and pharmacists, were assigned to systematically search for, review, and summarize emerging research for one or more novel therapeutics, including chloroquine, hydroxychloroquine, azithromycin, convalescent plasma, darunavir, lopinavir, ritonavir, tocilizumab, and remdesivir, on a weekly to biweekly basis. Both preprints and peer-reviewed publications were included in these searches. Manuscripts posted on Pubmed,medRxiv, bioRxiv, as well as journal websites, were included. Data from these summaries were then further extracted and organized into a systematic summary format by researchers with expertise in epidemiology and biostatistics.

##### Preprint

We systematically searched for studies of treatments for COVID-19. We searched the medRxiv, bioRxiv, arxiv, SSRN Electronic Journal, ChemRxiv, JMIR Preprints, Research Square databases with the following search terms: “COVID-19” or “SARS-CoV-2” and “clinical” and “treatment” and the name of the treatment. The treatments include chloroquine, hydroxychloroquine, azithromycin, carrimycin, convalescent plasma, ASC09, atazanavir, darunavir, danoprevir, lopinavir, ritonavir, stem cells, clazakizumab, olokizumab, sarilumab, siltuximab, sirukumab, tocilizumab, and remdesivir.

#### Eligibility Criteria

Eligibility was restricted to studies of treatments or drug-related human experimental studies for COVID-19. Studies were excluded with study type as vitro or animal testing. We also excluded the literature reviews and meta-analyses on COVID-19 treatments.

#### Selection process

Preprint titles and abstracts will be independently screened by three reviewers (A.B., A.P., O.W.). Based on the prespecified selection criteria, three reviewers independently identified studies. Disagreements were solved by discussion. Any unsettled conflict would be determined by a fourth reviewer (J.T.). The data collection forms were designed by the reviewers to extract the required data from the eligible studies. The reviewers assessed the extracted studies for duplication by comparing authors’ names, dates of publication or post, bioRxiv or medRxiv links of preprints, and population sizes.

#### Data extraction

For the survival cure model, confidence score calculation and case study, we extracted required data from the eligible studies, including study general information (title, authors’ names), publication/post date, population sizes, countries, single or multiple center studies, study types, number of arms, sizes of the arms, number of deaths in the arms, participants’ median ages, preliminary results or not, adjusted analysis or not, number of citations/usages as of September 03, 2020, and h-index of last author. Other data of interest included number of males, mortality rates, p-values of mortality rates, rates of cured patients, and symptoms.

### Statistical Analysis

In **Figure 2**, we illustrate the publication pipeline of a manuscript. A manuscript may be submitted directly to a journal without posting to a preprint server, after which it undergoes a review process by the journal that eventually leads to publication or non-publication. From this process, we only observe studies that have been published. Alternatively, an article may be posted to a preprint server either before submission to a journal or while it is under review. In this case, a preprint may be ‘published’, ‘not yet published’ (eg. are still undergoing review or have yet to be submitted), or ‘never published’ (eg. if they were rejected by the journal or if the authors do not intend to submit the manuscript for publication), though unlike direct submission to a journal, we can observe the preprints regardless of their publication status. Preprints that have not been published in a journal by the date of administrative censoring are either ‘not yet published’ or ‘never published’. Although we cannot directly observe which of these two categories the unpublished preprints fall into, we can model the probability that the manuscripts will be published by time t from the date of posting to the server.

**Figure 2.**
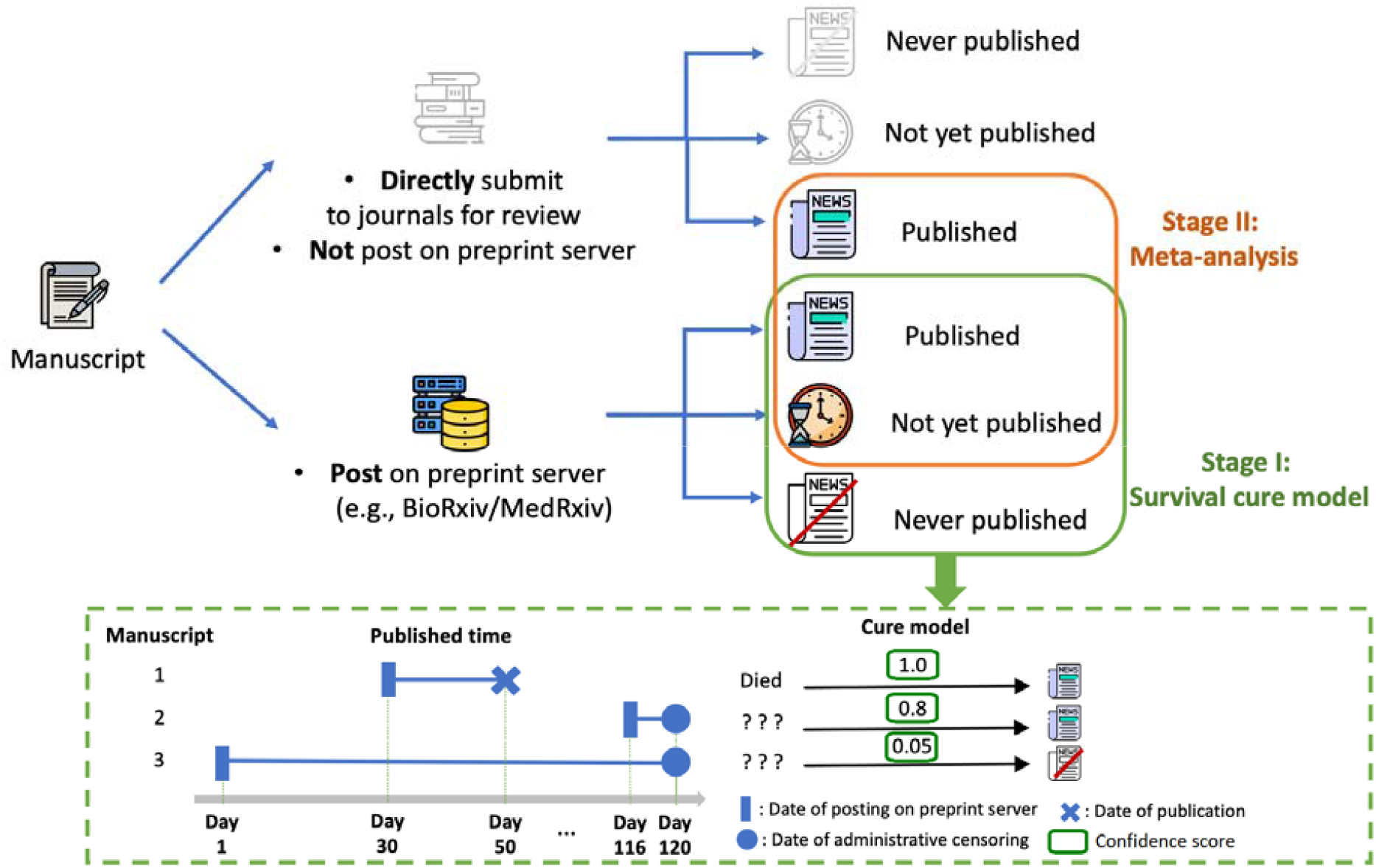
Publication pipeline of a manuscript. Events in grey are unobserved

To further illustrate the rationale behind our cure model, we consider three preprint manuscripts as an example in **Figure 2**. Preprint #1 is observed to be published 20 days after being posted to the server, and so it is assigned a weight (i.e., probability of being published, confidence score), of 1 in our meta-analysis, to represent the certainty with which we know it is published and should therefore incorporate the results in meta-analysis for evidence synthesis. Preprints #2 and #3 are administratively censored, thus we do not observe whether they will eventually be published or not. However, since preprint #2 was posted only 4 days prior to censoring, it has a higher likelihood of being published compared to preprint #3, which has been in circulation for 4 months. We would therefore assign preprint #2 greater weight relative to preprint #3 in our meta-analysis to reflect the higher ‘confidence’ we have that it will be published. For example, suppose 20% of the papers in medRxiv get published, then preprint #2 may have a 20% chance of getting published, while preprint #3 may have a much lower probability. These weights, or scores, may be modified when we additionally consider study-level characteristics that can impact the likelihood of publication, such as study type (e.g., RCT, cohort study, case reports), multi-center vs. single center studies, sample size, international vs. non-international study, etc. For example, a preprint reporting on an RCT with 1,000 patients will have a much higher chance of publication than the marginal probability of 20%. Motivated by this consideration, we propose the following two-stage procedure.

The proposed evidence synthesis method has two major components: calculation of confidence score and meta-analysis.

#### Step 1. Calculation of Confidence Score

The survival cure model^10–12^ was used to calculate the probability that a preprint will eventually be published. We refer this probability as the confidence score (Stage I analysis in **Figure 2**). For each preprint, we determined whether it had been published and obtained the time from posting to publication, or administrative censoring at 09/03/2020. Let T denote the time to publication, 1 − *π* (*z*) denote the probability that a manuscript will never be published depending on some study-level characteristics *z*, and *S*_*T*,0_(*t*|*x*) be the probability of *T* > *t* for those preprints that will be published depending on some study-level characteristics *x*. The mixture cure model assumes that the probability that a preprint has not been published by time *t* is

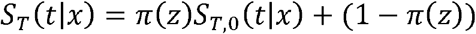

We use logistic regression with logit link for *π*(*z*) and Cox proportional hazard (PH) regression for *S*_*T*,0_ (t|*x*), i.e., *π* (*z*) = exp (*zβ*)/(1 + exp (*zβ*)) and *S*_*T*,0_ (*t*|*x*) = *S*_*T*,0_(*t*|0)^exp(*x β)*^, where *β* is the coefficient of the effects of *x*. The R package “*smcure*”^13^ was used to analyze the data. With this cure model, we can calculate the confidence scores for the preprints based on 10 extracted features, including study type (i.e., RCT, observational study, others), median age, sample size, single or multi-center study, if the result is preliminary, if the analysis is adjusted for confounding variables, country, citation counts as of 09/03/2020, number of pdfs downloaded on the preprint servers as of 09/03/2020, and h-index from Google scholar of the last author as of 09/03/2020.

#### Step 2. Evidence synthesis

After step 1, we synthesized both preprints and published articles in a meta-analysis. Suppose there are *n* total studies. For the i-th study, if it is not published yet, its probability of never being published is denoted by 1 − *π*_*i*_ from the survival cure model. The probability that it will eventually be published is denoted as *w*_*i*_ = *π*_*i*_. If the study is already published, we set *w*_*i*_ to be 1. Based on the *w*_*i*_, which characterizes the chance of publication for the i-th study, we propose the following *multiple imputation* procedure for evidence synthesis.

For each study, we impute the status of publication *u*_*i*_, where *u*_*i*_ = 1 indicates studies that will be published, and *u*_*i*_ = 0 indicates a study that will never be published, from a Bernoulli distribution with probability *w*_*i*_. We then conduct a random-effects meta-analysis using all the studies with *u*_*i*_ = 1. We can estimate an effect size, and standard error of the estimated effect size, as well as the heterogeneity variance *τ*^2^. By repeating the above imputation-estimation process multiple times, we can obtain a final estimate for the overall effect size by taking the average of the estimated effect sizes for all imputations, as well as a final estimate for the heterogeneity variance. In the case study, this imputation process was repeated 500 times.

## RESULTS

With the 158 preprints, which are related to COVID-19 treatments and collected till up to 09/03/2020, we fitted the multivariate survival cure model to calculate the confidence score by using10 covariates: study type (RCT or not), participants’ median age, sample size in log scale, single-center or multi-center, preliminary result or not, adjusted analysis or not, country (China, US, Europe, or others), h-index of last author, citation counts (standardized by length of being posted on preprint repos), PDF download counts in log scale.

In **Figure 3a**, we show the confidence score calculated by the cure model of all the 158 preprints. We grouped the confidence score based on the publication status. In the figure, the left column is for the unpublished preprints and the right one is for the published peer-reviewed preprints. At the end of 09/03/2020, 33 out of the 158 preprints have been published. For the unpublished preprints, the median of the confidence score is 0.013; for the published preprints, the median of the confidence score is 0.972, which is much larger than the former one. In addition, we evaluated the cure model by predicting the publication status of the 158 studies by 09/03/2020 (i.e., an in-sample prediction). The prediction by the predictive model is promising with an AUC value of 0.884 (**Figure 3b**)

**Figure 3.**
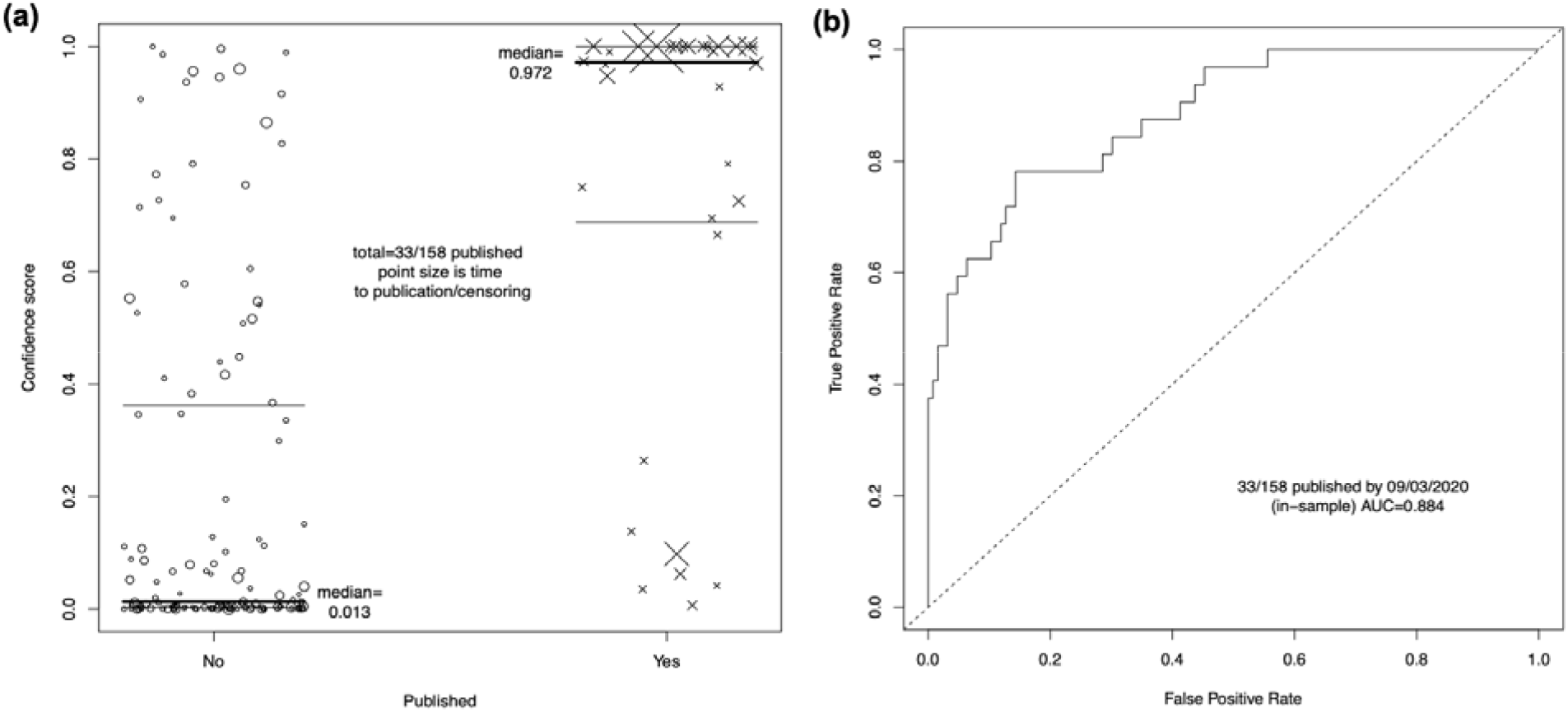
(a) Confidence score of the 158 preprints grouped by the publication status. The left column is the unpublished preprints and the right one is for the published studies. Each circle represents an unpublished study, and each cross is a published study. The solid thick horizontal lines are the medians of the scores, the thin lines above the median are the 75% quantile, and the ones below the median are the 25% quantile. (b) In-sample ROV curve and AUC value of the predictive model

## VALIDATION STUDY

To validate the proposed method, we manually collected 158 preprints from the preprint servers, and 19 papers which are peer-reviewed but not posted on preprint servers, as shown in the flow diagram (**Figure 4**). With the confidence score calculated by the cure model, a validation study has been conducted. With the mortality rate as the primary outcome, we compare the value of by including all 177 studies in a naive meta-regression with six covariates and the value of by using the multiple imputation method with the confidence scores. The result shows that the heterogeneity of the primary outcome is much more explained by using the latter method.

**Figure 4.**
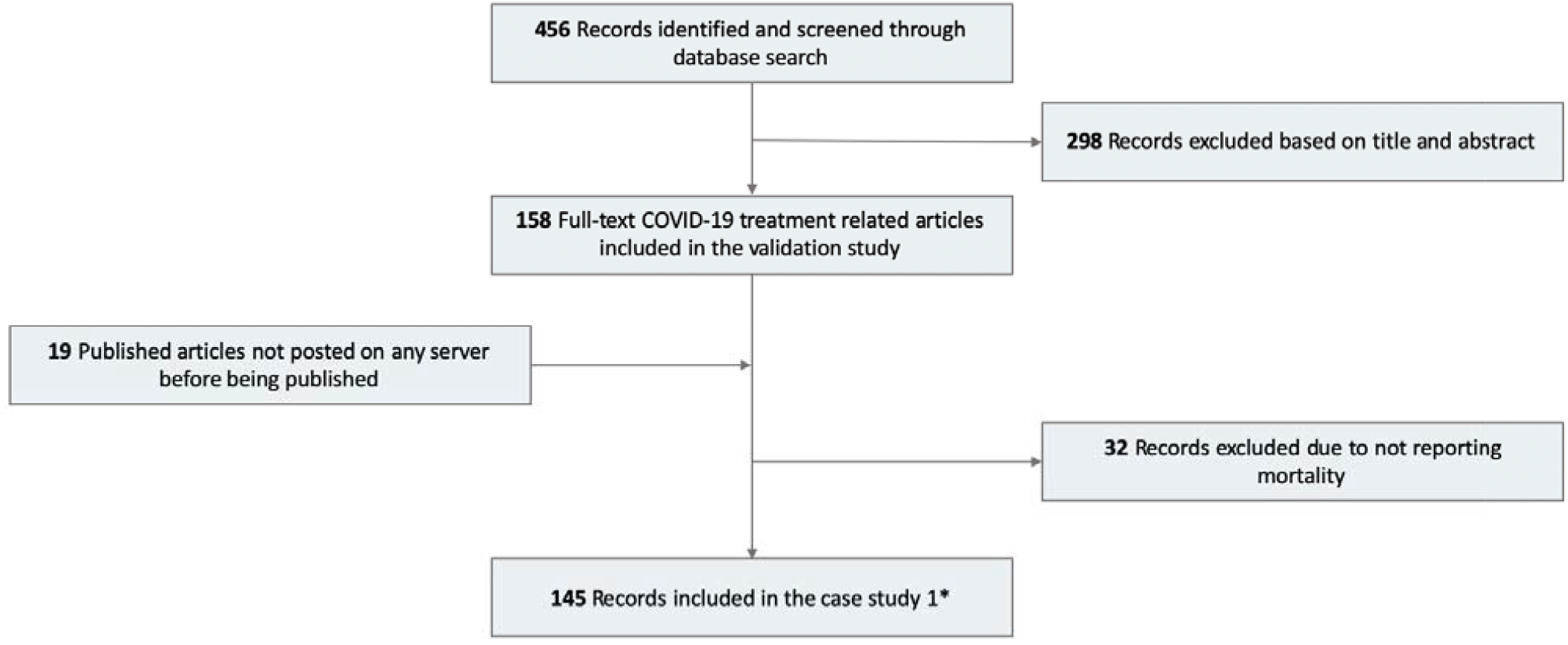
Flow diagram of systematic literature search for the meta-analysis

**Figure 5.**
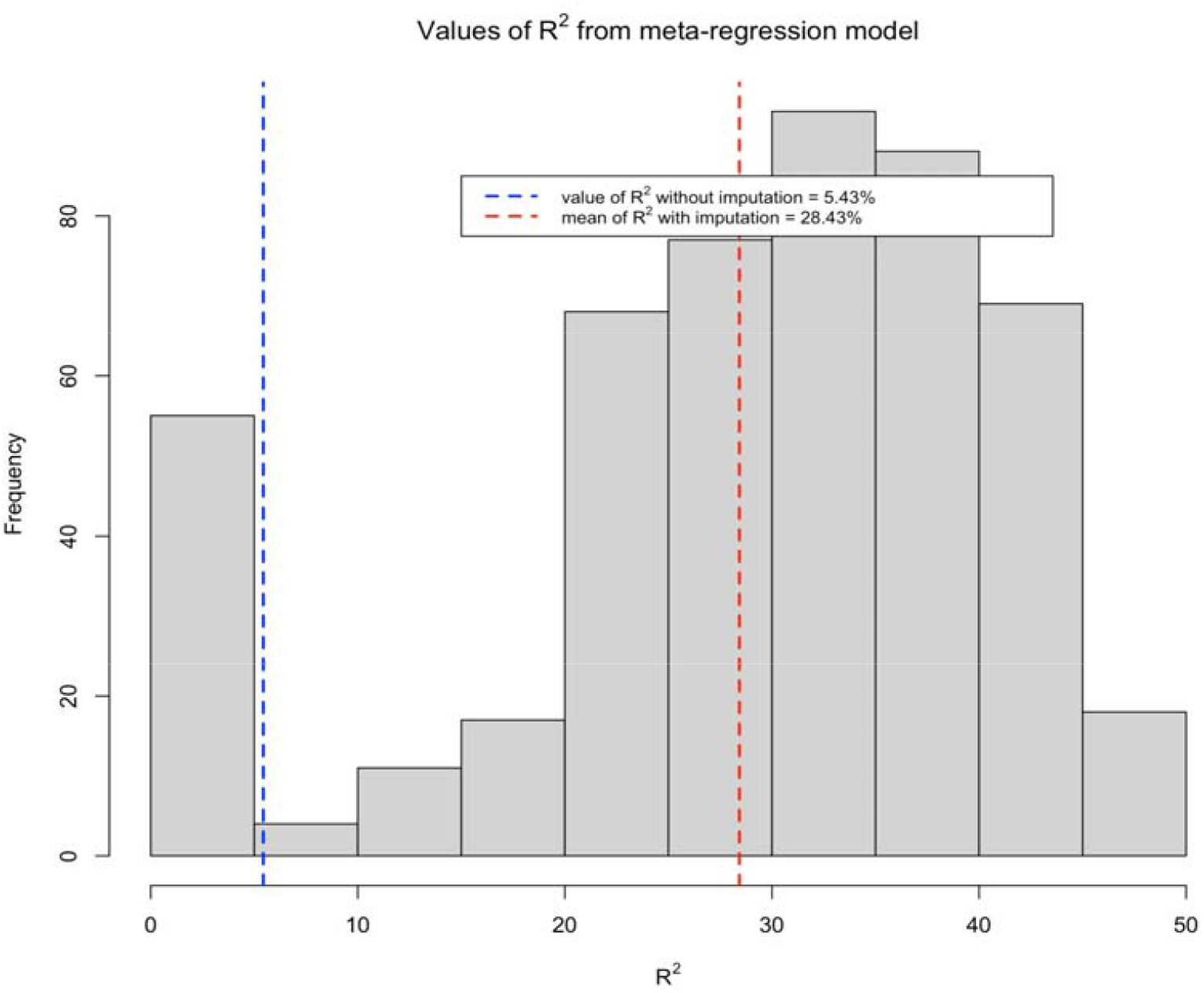
Histogram of values of 500-time imputation process. The blue dashed vertical line represents the value of without imputation process; the red dashed vertical line is the mean of through 500 times of imputation process

We have 177 studies including 19 studies that were peer-reviewed and not posted on any preprint servers and 158 preprints that were posted on the preprint servers. Among the 177 studies, there are 145 studies that have mortality rates reported, which is the primary outcome we use to validate the proposed method. These 145 studies include 19 peer-reviewed but not posted on preprint servers and 126 preprints.

First, we conducted a basic meta-analysis with the mortality rate as the primary outcome. However, the mortality rates reported by the studies for COVID-19 are highly heterogeneous. Measures of heterogeneity (= 97.0%, Cochrane’s Q-test p-value < 0.01) suggest evidence of statistical heterogeneity. The excessive heterogeneity can be caused by different study-level characteristics and study designs. For example, some studies reported the 14-day mortality, but others reported 30-day or 28-day mortality. The severity conditions of the patients who were recruited/included into the studies also varies case by case. Thus, we use meta-regression by including 8 covariates into the model: indicator of preprint or not, mean age, country, single or multiple center study, study type, drug class, sample size, and number of citations. The value of for the meta-regression with 145 studies is 5.43%, which means the six covariates included in the regression model are able to explain 5.43% of the heterogeneity in the mortality rates. With the imputation-estimation process, we expected more heterogeneity can be explained by the regression model.

Following the imputation process, we repeat it for 500 times by weighing the finally published papers with 1 and the not published papers with the confidence scores from the cure model. The following figure present the histogram of the values for 500 times. The x-axis is the values of and the y-axis represents the frequency. The dotted blue line is the value of for meta-regression without imputation process (5.43%) and the dotted red link is the mean value of for meta-regression with the imputation process (28.43%). Among the 500 times of imputation processes, 88.8% of them have larger value of than 5.43%.

## DISCUSSION

In this paper, we proposed a novel method that provides a strategy to include preprints in the systematic reviews and meta-analyses. In particular, we used the survival cure model to predict the probability of preprint to be published and termed this probability as the confidence score. The confidence score was then used as the weight of a preprint in the multiple imputation process in a meta-analysis study. To evaluate the proposed method, we conducted systematic reviews on the preprints, identified 158 preprints based on our inclusion criteria, and calculate the confidence scores. We further used a case study with 145 records to validate the confidence score. The proposed approach provides the solution to the incorporation of preprints in evidence synthesis. With the confidence score which accounts for the publication probability, the preprints can be appropriately integrated into meta-analyses.

The proposed method by including preprints in evidence synthesis is able to assist the living systematic reviews process. Despite the promise of producing high-quality and timely evidence syntheses, living systematic reviews and cumulative meta-analyses require extraordinary long-term commitments from large research teams; for example, Siemieniuk et al.^16^ published and maintain a living systematic review of network meta-analyses, comparing 23 treatments for COVID-19, which involved effort from a team of 58 researchers across 33 institutions. It is well understood that active evidence monitoring is time-consuming and labor-intensive. Even after the initial living systematic review is published, it requires continued effort to maintain and update the results online. To address these logistical challenges, our proposed framework reduces the efforts without requiring long-term commitments and provides high quality evidence synthesis by including preprints.

On the other hand, the proposed framework can be extended to a living systematic review if there are sufficient resources. With further investigation of treatments for COVID-19, a growing amount of knowledge may be offered as preprints over time. As more evidence is collected through living systematic reviews by continuous and active monitoring, a data-driven cure model can enable the incorporation of preprints in this process to sustain and strengthen the validity of results. The proposed framework can be particularly extended to cumulative meta-analyses (which are meta-analyses that are updated every time new evidence appears, to allow the study of temporal trends of intervention/treatment effects^17^).

In terms of human labor, the limitation of including preprints in the proposed method is that the searching for the preprints would be intensive. To better train the cure model, the more preprints we have the more accurate prediction we’ll get. Thus, we search across several preprint servers including medRxiv, bioRxiv, and arxiv for the COVID-19 related articles with the top 5 therapies from COVID-19 Clinical Trials Explorer^18^. At this stage, the strategy of being inclusive would lead to a large number of samples to train the cure model. Although the search of preprints is labor-intensive, this task only requires the labor from people who have less domain expertise. This is because, at this stage, no screenings or annotations would be executed. In addition, the efforts that have been made so far would contribute to saving more labor in the future. The current data which are extracted manually can be used as a training dataset to develop a semi-automated NLP tool, which will automate the data extraction process in the future. Once we have our cure model with good performance, the confidence scores predicted by the model can provide convincing evidence for the researchers to review the preprints. The domain experts can choose a subset from the preprints to review based on the confidence score instead of reviewing all of them. In other words, the cure model does save the labor from domain experts in the systematic review process.

Currently, we only focus on the searching and prediction for COVID-19 related articles. The proposed method also has broad applicability to other research topics related to COVID-19 (such as the effectiveness of vaccines in prevention) and other quickly evolving fields outside of COVID-19, such as cardiology and oncology. There is a chance that we can borrow and fine-tune the model. However, we need to be careful when adjusting the model for the diseases by considering particular features of the diseases. For the existing diseases which have been studied for several years or decades, the consensus of the treatments or interventions for the diseases could have been reached through numbers of publication. In this case, we might not need to include the preprints.

## Data Availability

Data for this study are unavailable publicly and may be made available to the editors upon reasonable request.

